# Genetic and other omics-based information in the most-cited recent clinical trials

**DOI:** 10.1101/2024.10.21.24315878

**Authors:** Luigi Russo, Leonardo M. Siena, Sara Farina, Roberta Pastorino, Stefania Boccia, John P.A. Ioannidis

## Abstract

**Importance:** Genetics and other -omics’ technologies have long been proposed for medical research use.

**Objective:** To assess how genetics and other -omics information’ are used in the most cited recent clinical trials, and to evaluate industry involvement and transparency patterns.

**Design, Setting, Participants:** Meta-research evaluation using a previously constructed database of the 600 most cited clinical trials published from 2019 to 2022.

**Main Outcome(s) and Measure(s):** Utilization of genetic or other -omics information in the trial design, analysis, and results; industry involvement and transparency.

**Results:** 132 (22%) trials used genetic or other -omics information, predominantly for detection of cancer mutations (n=101). Utilization included eligibility criteria (n=59), subgroup analysis (n=82), and stratification factor in randomization (n=14). Authors addressed the relevance in the conclusions in 82 studies (62%). 102 studies (77%) provided data availability statements and 6 had data already available. Most studies had industry funding (n=111 [84.0%]). Oncology trials were more likely to be industry-funded (90.1% vs 64.5%, p=0.001), to have industry-affiliated analysts (43.6% vs 22.6%, p=0.036) and to favor industry-sponsored interventions (83.2% vs 58.1% p=0.004). When compared to other trials, genetic and other -omics trials were more likely to be funded by industry (84% vs 63.9%, p<0.001) and tended to be less likely to have full protocols (p=0.018) and statistical plans (p=0.04) available.

**Conclusions and Relevance:** Our study highlights the current underutilization of genetic and other-omics technologies beyond testing for cancer mutations. Industry involvement in these trials appears to be more substantial and transparency is more limited, raising concerns about potential bias.

**Key Points:** *Question:* How are genetics and other -omics information used in highly impactful clinical trials, and what is the involvement of the industry in their development?

*Findings:* 22% of the trials employed genetic or -omics information, with the vast majority being oncology-related and focusing on testing for mutations. Industry was more heavily involved in the funding and in the design of these trials, in particular for oncology trials.

*Meaning:* The use of genetics and -omics is limited and needs to become more common in non-oncology trials. Industry involvement is intense in this context, highlighting the need for improvements in transparency and independence.

## Introduction

Since the genomic revolution in the early 2000s and the launch of the precision medicine initiative in 2015^1^, the use of genetic and genomic technologies has significantly expanded within clinical studies. Advances in genomic sequencing technologies have targeted therapies in oncology ^2,3^. Personalized medicine is also being applied in other fields, such as cardiology, where its use is being explored for diverse conditions, including coronary artery disease, arrhythmias, and cardiomyopathies^4^. As research continues to evolve, one may anticipate even broader applications of these technologies across various medical disciplines.

This rise of personalized healthcare may reshape the design and analysis of clinical trials ^5,6^. Genetic and other -omics data can be utilized in trial design, including participant selection, group stratification, outcome assessment, and the examination of differences across participants with various genetic and/or -omics profiles. Understanding the frequency and extent to which influential clinical trials utilize genetic and/or -omics information in these ways would be highly valuable. A preceding study created a database of the top 600 most-cited clinical trials published from 2019 to 2022 across medicine, highlighting the significant role of industry involvement in these studies and identifying areas where transparency could be improved ^7^. Building on this database, we aimed to investigate the use and impact of genetic and other -omics technologies within these influential trials. Specifically, the current study aimed to assess how frequently genetics and -omics information was used in the design and analysis of trials, in detecting significant differences in subgroups, and in influencing the authors’ conclusions in the article abstracts. Additionally, we evaluated differences in transparency and funding models between trials that use genetic and -omics information and those that do not.

## Methods

### Sample of Eligible Trials

The analysis was based on the 600 most-cited clinical trials published between 2019 and 2022, as identified in a previous cross-sectional study ^7^. Trials were defined as research studies where human participants were prospectively assigned to one or more interventions to evaluate their effects on health-related outcomes. Only trials published in English were included, and articles reporting on more than one trial were excluded. Both randomized and nonrandomized trials were eligible. For the current study, we focused specifically on the subset of trials that employed genetic or - omics characterization of participants (including genotyping, metabolomics, proteomics, transcriptomics, or any other -omics platforms) at any stage of the trial’s design, analysis, and/or interpretation. Trials that used immunochemistry to identify biomarkers or those that employed methods other than genetic or -omics testing were excluded. Secondary or additional publications employing such measurements but not included among the 600 most-cited main articles were not considered.

The protocol of this study has been registered on the Open Science Framework before data extraction ^8^.The reporting of this study follows the Strengthening the Reporting of Observational Studies in Epidemiology (STROBE) reporting guideline for observational studies^9^.

### Search Strategy and Screening

The full texts of the 600 articles from the previous project were available for review. Each article was independently assessed by two researchers, with a third reviewer resolving any discrepancies. Articles identified as employing genetic or -omics characterization of participants in any aspect of the trial’s design, analysis, and/or interpretation were then subjected to detailed data extraction. A full list of included studies is provided in Supplement 1.

### Data Extraction

From each included study, two independent investigators manually retrieved previously extracted information or undertook extraction as necessary, with a third reviewer arbitrating disagreements. The following general information was collected: title, first author, DOI, journal of publication, year of publication, and trial location. For study design, we recorded the number of arms, the number of participants in each arm, and the total number of participants. We categorized the medical specialty (e.g., radiology, cardiology, pulmonology) and specific disease (e.g., diabetes, Parkinson’s disease) under investigation, noting the use of novel trial designs (e.g., enrichment, randomize-all, adaptive design, umbrella, basket) ^5^. Regarding genetic or -omics characterization, we documented the type of -omics in study, the type of measurements made, their role in eligibility criteria (inclusion and/or exclusion), their use as a stratification factor in randomization, their role in determining treatment changes during follow-up (adaptive design), and any other application in the trial design. We also recorded the use of genetic or -omics data in subgroup or stratified analyses of outcomes, the specific outcomes assessed, whether any differences observed were statistically significant beyond chance, (evaluated with p<0.05 or p<0.005), and whether the authors mentioned these findings in the abstract.

Data on industry involvement and transparency features were retrieved from the previous dataset. This included information on author affiliation, funding sources, whether the analysis was conducted by industry analysts, and whether the trial conclusions favored an industry-sponsored intervention. Transparency features evaluated included the presence of a data availability statement, the declared availability of data, the type of data available, access conditions, protocol and statistical analysis plan availability, and whether analysis code was shared.

A pilot test involving four full texts was conducted to validate the extraction process and ensure consistency in data collection.

### Main Outcomes

Primary outcomes were the proportions of trials that used genetic and/or other -omics information in the design of the trials (including inclusion and exclusion criteria, stratification, and adaptive treatment decisions); and in trial analysis (subgroup or stratified analyses based on genetic or - omics data); the frequency of results of genetic or other omics-based analyses with statistically significant differences (at p<0.05 and p<0.005 levels); and the influence of genetic or -omics data on the conclusions drawn in the article abstracts. Given that oncology trials were found to be the large majority of those eligible, after the protocol had been posted we also added analyses that separated oncology and other trials and compared the two sets.

We also analyzed funding and transparency patterns to explore whether medical specialty, type of study, type of funding, author affiliation, analyst affiliation, data availability, protocol availability, statistical plan availability and result direction differed between trials that used genetic or -omics data and those that did not.

### Statistical Analysis

Descriptive statistics of trial characteristics and main outcomes were reported as counts, percentages, mean (or median) with standard deviation (SD) (or interquartile range (IQR), or range), along with corresponding 95% CIs, as appropriate. We used Pearson’s χ² or Fisher’s exact tests to assess whether disease type, study type, industry funding, author affiliation, analyst affiliation, data availability, protocol availability, statistical plan availability, and result favorability toward sponsored interventions differed between trials utilizing genetic and -omics data versus those that did not. A 2-sided P < .005 was considered statistically significant, and P < .05 was considered suggestive ^10^. All analyses were performed using Stata software version 17.0 (StataCorp).

## Results

Out of the 600 most highly cited articles, 132 (22%) used genetic and/or other -omics information and were eligible. These trials had a median (IQR) sample size of 208 (61-433) participants. As shown in Table 1, two thirds of the clinical trials involved collaboration among multiple countries. Among the single-country trials, the United States was the most common location. Two-thirds of the studies were randomized, while the remaining one-third were nonrandomized single-group studies. The vast majority focused on oncological conditions (76.5%).

**Table 1.** Characteristics of the included trials (N = 132)

| **Variable** | **Studies No.** | **%** |
| --- | --- | --- |
| **Year** |  |  |
| 2019 | 78 | 59.1% |
| 2020 | 49 | 37.1% |
| 2021 | 5 | 3.8% |
| 2022 | 0 | 0.0% |
| **Type of study** |  |  |
| Randomized | 81 | 61.4% |
| Nonrandomized single arm | 51 | 38.6% |
| Nonrandomized controlled | 0 | 0.0% |
| **Intervention comparison** | **N** | **%** |
| Superiority | 71 | 53.7% |
| Non inferiority | 2 | 1.6% |
| Equivalence | 0 | 0.0% |
| Not applicable | 59 | 44.7% |
| **Location** |  |  |
| International | 84 | 63.6% |
| United States | 21 | 15.9% |
| United Kingdom | 4 | 3.0% |
| China | 3 | 2.3% |
| France | 3 | 2.3% |
| The Netherlands | 2 | 1.5% |
| Other | 15 | 11.4% |
| **Disease in study** |  |  |
| Oncology | 101 | 76.5% |
| *Non-small cell lung cancer (NSCLC)* | 28 | 21.2% |
| *Hematologic malignancies* | 20 | 15.2% |
| *Colorectal cancer* | 7 | 5.3% |
| *Prostate cancer* | 6 | 4.5% |
| *Ovarian cancer* | 5 | 3.8% |
| *Breast cancer* | 5 | 3.8% |
| *Endometrial cancer* | 3 | 2.3% |
| *Others tumors* | 27 | 20.5% |
| Gastroenterology | 4 | 3.0% |
| Cardiology | 2 | 1.5% |
| Infectious diseases | 1 | 0.8% |
| Other diseases | 24 | 18.2% |

### Characteristics of genetic or other -omics tests used

Genetics and genomics applications were the most frequently used, appearing together in 116 studies, with genetics and genomics alone used in 60.6% and 32.6% of the studies, respectively (Table 2). The primary use of these applications was the detection of specific genetic mutations (n=101, 76.5% of the studies). Among these, 87 studies focused on somatic mutations, while 14 studies examined germline mutations. 36 studies (27%) employed multiple applications (data not shown).

**Table 2.** Characteristics of genetic or other -omics tests used in the included trials (N= 132)

| Studies, No. (%) |  |  |  |  |
| --- | --- | --- | --- | --- |
| Variables | All trials<br>(n=132) | Oncology<br>trials (n=101) | Non-oncology<br>trials (n=31) | p-value |
| **Type of –omics<sup>a</sup>** |  |  |  |  |
| Genetics | 80 (60.6%) | 69 (68.3%) | 11 (35.5%) | 0.001 |
| Genomics | 43 (32.6%) | 35 (34.7%) | 8 (25.8%) | 0.350 |
| Transcriptomics | 27 (20.5%) | 20 (19.8%) | 7 (22.6%) | 0.737 |
| Proteomics | 14 (10.6%) | 7 (6.9%) | 7 (22.6%) | 0.013 |
| Metagenomics | 11 (8.3%) | 2 (2.0%) | 9 (29.0%) | <0.001 |
| Metabolomics | 11 (8.3%) | 2 (2.0%) | 9 (29.0%) | <0.001 |
| **Type of measurements<sup>a</sup>** |  |  |  |  |
| Genetic mutation | 101 (76.5%) | 87 (86.1%) | 14 (45.2%) | <0.001 |
| *Somatic* | 87 (86.1%) | 84 (83.2%) | 3 (9.7%) | <0.001 |
| *Germline* | 14 (13.9%) | 3 (3.0%) | 11 (35.5%) | <0.001 |
| Gene expression | 20 (15.2%) | 14 (13.9%) | 6 (19.4%) | 0.456 |
| Microbiome | 9 (6.8%) | 1 (1.0%) | 8 (25.8%) | <0.001 |
| 16S rRNA sequencing | 6 (4.5%) | 0 (0.0%) | 6 (19.4%) | <0.001 |
| Tumor mutational burden | 6 (4.5%) | 6 (5.9%) | 0 (0.0%) | 0.165 |
| Metabolite expression | 6 (4.5%) | 1 (1.0%) | 5 (16.1%) | <0.001 |
| CAR T | 4 (3.0%) | 4 (4.0%) | 0 (0.0%) | 0.261 |
| Transcriptome | 2 (1.5%) | 1 (1.0%) | 1 (3.2%) | 0.373 |
| Others | 8 (6.1%) | 4 (4.0%) | 4 (12.9%) | 0.068 |
| **Test used as eligibility criterion** |  |  |  |  |
| Yes | 59 (44.7%) | 48 (47.5%) | 11 (35.5%) | 0.238 |
| No | 73 (55.3%) | 53 (52.5%) | 20 (64.5%) |  |
| **Test used as stratification factor in randomization** |  |  |  |  |
| Yes | 14 (10.6%) | 11 (10.9%) | 3 (9.7%) | 0.848 |
| No | 118 (89.4%) | 90 (89.1%) | 28 (90.3%) |  |
| **Test used for subgroups or stratified analysis** |  |  |  |  |
| Yes | 82 (62.1%) | 69 (68.3%) | 13 (41.9%) | 0.008 |
| No | 50 (37.9%) | 32 (31.7%) | 18 (58.1%) |  |
| **Results of the subgroups or stratified analysis** |  |  |  |  |
| *Considering the trials in which the test is used for subgroup analysis (N = 82)* |  |  |  |  |
| Statistically significant <sup>b</sup> | 50 (61.0%) | 43 (52.4%) | 7 (8.5%) | 0.566 |
| *Somatic mutation* | 40 (48.8%) | 39 (47.6%) | 1 (1.2%) | <0.001 |
| *Germinal mutation* | 3 (3.7%) | 2 (2.4%) | 1 (1.2%) | 0.320 |
| *Others measurement (i.e. metabolite expression profile)* | 7 (8.5%) | 2 (2.4%) | 5 (6.1%) | <0.001 |
| Not statistically significant <sup>^</sup> | 32 (39.0%) | 26 (31.7%) | 6 (7.3%) |  |
| *Somatic mutation* | 24 (29.3%) | 22 (26.8%) | 2 (2.4%) | 0.009 |
| *Germinal mutation* | 2 (2.4%) | 0 (0.0%) | 2 (2.4%) | 0.002 |
| *Others measurement (i.e. metabolite expression profile)* | 6 (7.3%) | 4 (4.9%) | 2 (2.4%) | 0.310 |
| **Outcomes assessed in trials conducting subgroups or stratified analyses according to genetics and other -omics test<sup>c</sup>** |  |  |  |  |
| *Considering the trials in which the test is used for subgroup analysis (N = 82)* |  |  |  |  |
| Progression Free Survival | 31 (37.8%) | 31 (37.8%) | 0 (0.0%) | 0.002 |
| Overall Survival | 22 (26.8%) | 22 (26.8%) | 0 (0.0%) | 0.017 |
| Response to treatment | 27 (32.9%) | 25 (30.5%) | 2 (2.4%) | 0.142 |
| Overall survival and Progression free survival | 8 (9.8%) | 8 (9.8%) | 0 (0.0%) | 0.196 |
| Death | 6 (7.3%) | 5 (6.1%) | 1 (1.2%) | 0.955 |
| Remission | 5 (6.1%) | 4 (4.9%) | 1 (1.2%) | 0.793 |
| Recurrence | 6 (7.3%) | 6 (7.3%) | 0 (0.0%) | 0.269 |
| Microbiome changes | 3 (3.7%) | 0 (0.0%) | 3 (3.7%) | <0.001 |
| LDL – C<sup>d</sup> change | 2 (2.4%) | 0 (0.0%) | 2 (2.4%) | 0.001 |
| **Conclusions include consideration of genetic and -omics information** |  |  |  |  |
| Yes | 82 (62.1%) | 64 (63.4%) | 18 (58.1%) | 0.595 |
| No | 50 (37.9%) | 37 (36.6%) | 13 (41.9%) |  |

<sup>a</sup>same studies employed multiple measurements; <sup>b</sup>for the statistical significance definition see methods; <sup>c</sup>multiple outcomes were evaluated by the same studies; <sup>d</sup>low-density lipoproteins cholesterol;

Fifty-nine trials (44.7%) used genetics and other-omics as eligibility criteria, while in 14 studies (10.6%), a genetic or -omics measurement was used as stratification factor in participants’ randomization (Table 2). Among the 82 trials (62.9%) in which subgroup or stratified analysis was conducted according to the genetic or other –omics test, in 60.2% (50/82) it showed a significant result. Moreover, genetic or other -omics information was taken into consideration in the conclusions of 62.1% of the trials (Table 2).

Oncology trials were more likely to perform a genetic measurement (68.3% vs 35.5%, p=0.001), in particular genetic testing for somatic mutation (p<0.001), and had a trend to use more frequently such measurement for a subgroup or stratified analysis (68.3% vs 41.9%, p=0.008) when compared to non-oncology trials. Non-oncology trials were more likely to employ metagenomics and metabolomics measurements (29% vs 2%, p<0.001), to perform microbiome analysis (p<0.001), 16S rRNA sequencing (p<0.001) and metabolite expression evaluation (p<0.001). Furthermore, non-oncology trials had a higher chance of performing genetic testing for germline mutation (3.0% vs 35.5%, p<0.001).

### Funding and Transparency Features

Sixty percent of the trials were exclusively funded by industry, with 111 trials (84.0%) having industry at least as a co-founder; among those the majority (91.8%) concluded in favor of the intervention. Regarding transparency, 102 trials (77.3%) provided a data availability statement (Table 3), with most indicating an intention to share data, and in 47 trials (54.0%), this was as anonymized individual participant data. However, data were already available at the time of extraction in only six studies (6.8%). Furthermore, the 75% of the trials had a full protocol available, the 67.4% had accessible statistical analysis plans, and six studies (4.5%) explicitly mentioned that they would share the statistical analysis code.

**Table 3.**
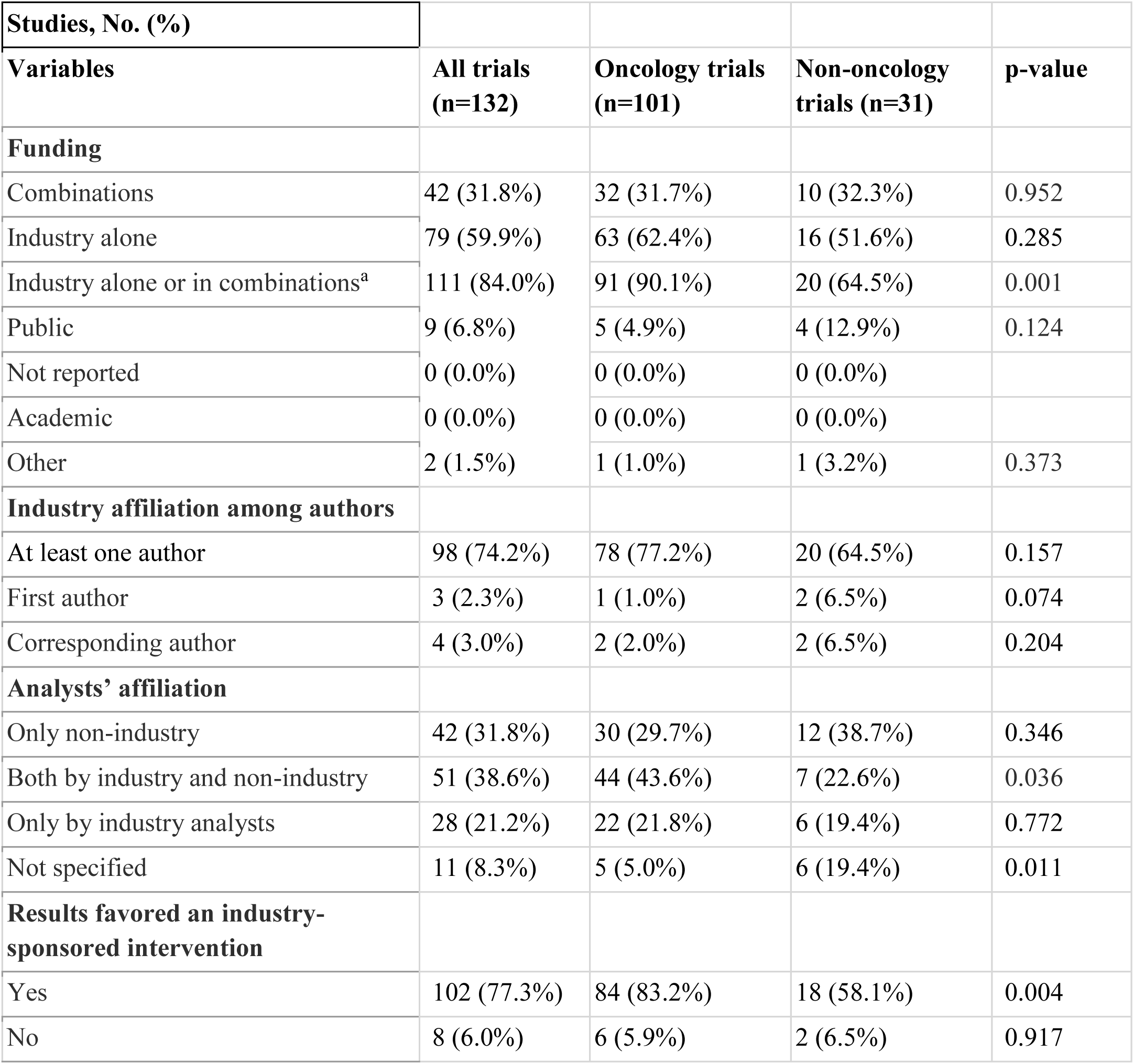
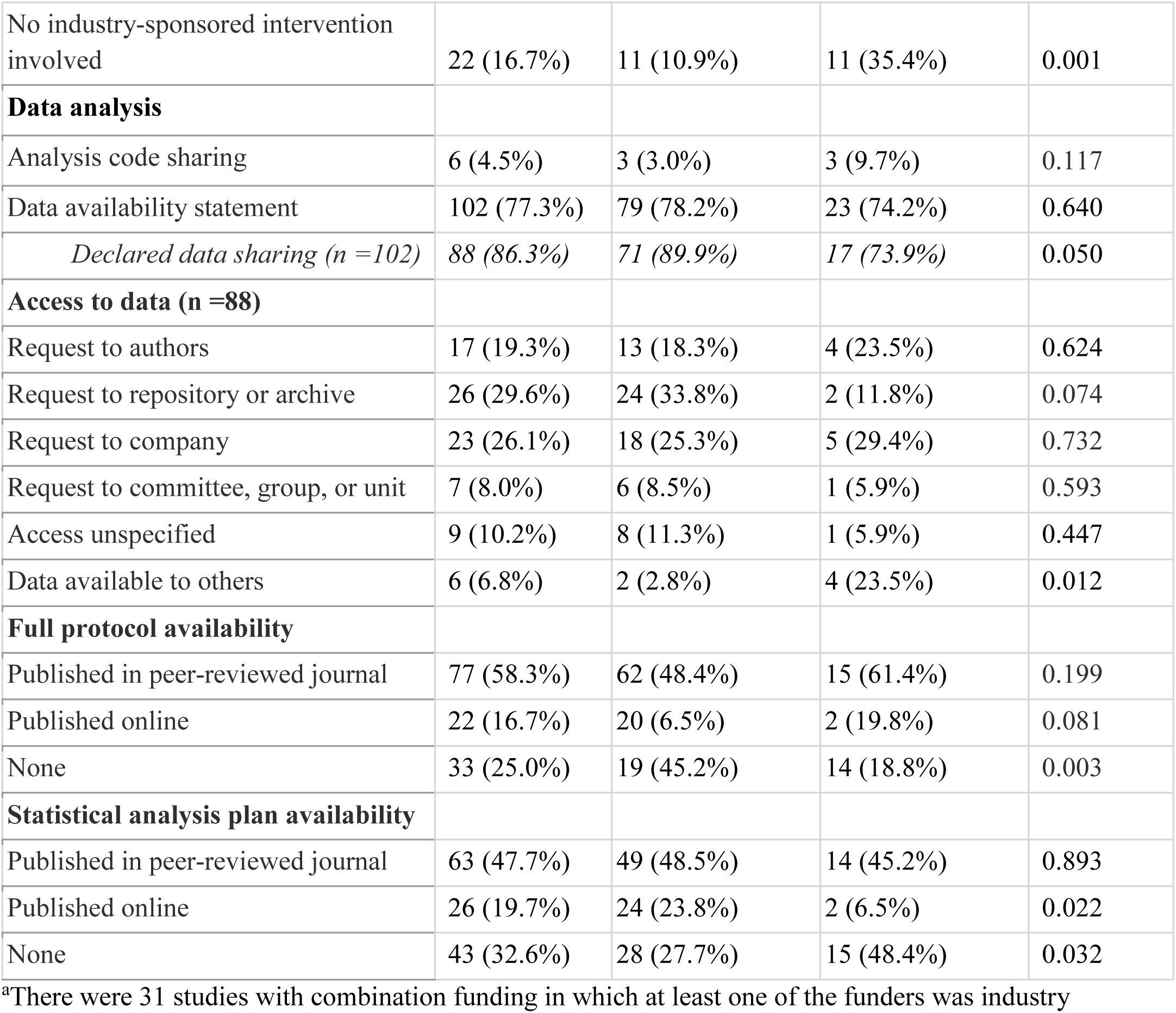
Transparency and Funding Features in genetic or -omics trials (N= 132)

| **Studies, No. (%)** |  |  |  |  |
| --- | --- | --- | --- | --- |
| **Variables** | **All trials (n=132)** | **Oncology trials (n=101)** | **Non-oncology trials (n=31)** | **p-value** |
| **Funding** |  |  |  |  |
| Combinations | 42 (31.8%) | 32 (31.7%) | 10 (32.3%) | 0.952 |
| Industry alone | 79 (59.9%) | 63 (62.4%) | 16 (51.6%) | 0.285 |
| Industry alone or in combinations<sup>a</sup> | 111 (84.0%) | 91 (90.1%) | 20 (64.5%) | 0.001 |
| Public | 9 (6.8%) | 5 (4.9%) | 4 (12.9%) | 0.124 |
| Not reported | 0 (0.0%) | 0 (0.0%) | 0 (0.0%) |  |
| Academic | 0 (0.0%) | 0 (0.0%) | 0 (0.0%) |  |
| Other | 2 (1.5%) | 1 (1.0%) | 1 (3.2%) | 0.373 |
| **Industry affiliation among authors** |  |  |  |  |
| At least one author | 98 (74.2%) | 78 (77.2%) | 20 (64.5%) | 0.157 |
| First author | 3 (2.3%) | 1 (1.0%) | 2 (6.5%) | 0.074 |
| Corresponding author | 4 (3.0%) | 2 (2.0%) | 2 (6.5%) | 0.204 |
| **Analysts' affiliation** |  |  |  |  |
| Only non-industry | 42 (31.8%) | 30 (29.7%) | 12 (38.7%) | 0.346 |
| Both by industry and non-industry | 51 (38.6%) | 44 (43.6%) | 7 (22.6%) | 0.036 |
| Only by industry analysts | 28 (21.2%) | 22 (21.8%) | 6 (19.4%) | 0.772 |
| Not specified | 11 (8.3%) | 5 (5.0%) | 6 (19.4%) | 0.011 |
| **Results favored an industry-sponsored intervention** |  |  |  |  |
| Yes | 102 (77.3%) | 84 (83.2%) | 18 (58.1%) | 0.004 |
| No | 8 (6.0%) | 6 (5.9%) | 2 (6.5%) | 0.917 |
| No industry-sponsored intervention involved | 22 (16.7%) | 11 (10.9%) | 11 (35.4%) | 0.001 |
| **Data analysis** |  |  |  |  |
| Analysis code sharing | 6 (4.5%) | 3 (3.0%) | 3 (9.7%) | 0.117 |
| Data availability statement | 102 (77.3%) | 79 (78.2%) | 23 (74.2%) | 0.640 |
| *Declared data sharing (n =102)* | 88 (86.3%) | 71 (89.9%) | 17 (73.9%) | 0.050 |
| **Access to data (n =88)** |  |  |  |  |
| Request to authors | 17 (19.3%) | 13 (18.3%) | 4 (23.5%) | 0.624 |
| Request to repository or archive | 26 (29.6%) | 24 (33.8%) | 2 (11.8%) | 0.074 |
| Request to company | 23 (26.1%) | 18 (25.3%) | 5 (29.4%) | 0.732 |
| Request to committee, group, or unit | 7 (8.0%) | 6 (8.5%) | 1 (5.9%) | 0.593 |
| Access unspecified | 9 (10.2%) | 8 (11.3%) | 1 (5.9%) | 0.447 |
| Data available to others | 6 (6.8%) | 2 (2.8%) | 4 (23.5%) | 0.012 |
| **Full protocol availability** |  |  |  |  |
| Published in peer-reviewed journal | 77 (58.3%) | 62 (48.4%) | 15 (61.4%) | 0.199 |
| Published online | 22 (16.7%) | 20 (6.5%) | 2 (19.8%) | 0.081 |
| None | 33 (25.0%) | 19 (45.2%) | 14 (18.8%) | 0.003 |
| **Statistical analysis plan availability** |  |  |  |  |
| Published in peer-reviewed journal | 63 (47.7%) | 49 (48.5%) | 14 (45.2%) | 0.893 |
| Published online | 26 (19.7%) | 24 (23.8%) | 2 (6.5%) | 0.022 |
| None | 43 (32.6%) | 28 (27.7%) | 15 (48.4%) | 0.032 |

<sup>a</sup>There were 31 studies with combination funding in which at least one of the funders was industry

Oncology trials were more likely than non-oncology trials to be funded by the industry (90.1% vs 64.5%, p=0.001), to have an analyst affiliated to the industry (43.6% vs 22.6%, p=0.036), having results favoring an industry-sponsored intervention (83.2% vs 58.1% p=0.004) and not having full protocol availability (45.2% vs 18.8%, p=0.003). Non-oncology trials tended to be more prone to make data available to others (23.5% vs 2.8%, p=0.012), and were more likely to not involve an industry sponsored intervention (35.4% vs 10.9%, p=0.001), but tended to be less likely to publish the statistical analysis plan online (6.5% vs 23.8%, p=0.022) or in general (p=0.032).

### Funding and transparency differences between genetic or -omics trials *vs* other trials

As shown in Table 4, genetic and other -omics trials tended to be more likely to be solely funded by the industry (59.9% vs. 47.4%, p=0.013), or were more likely to have any industry funding (84% vs 63.9%, p<0.001) while the authors were more frequently affiliated with the industry (74.2% vs. 54.7%, p<0.0001), compared to those in non-genetic trials. For trials funded or co-funded by industry, genetic and other -omics trials tended to be more likely to favor the sponsor compared to other trials (77.3% vs. 56%, p<0.001). Genetic/-omics trials tended to be more likely to not have full protocols available (25.0% vs. 16%, p=0.018) and to not have the statistical analysis plan available (32.6% vs. 23.7%, p=0.04) compared to other trials.

**Table 4.** Differences in funding and transparency in genetic or -omics trials vs other trials.

| **Studies, No. (%)** |  |  |  |
| --- | --- | --- | --- |
| **Variables** | **Genetic or -omics trials** | **Other trials<sup>a</sup>** | **p-value** |
| **Funding** |  |  |  |
| Combinations | 42 (31.8%) | 122 (26.1%) | 0.190 |
| Only Industry | 79 (59.9%) | 223 (47.7%) | 0.013 |
| Industry alone or in combination<sup>b</sup> | 111 (84.0%) | 299 (63.9%) | <0.001 |
| Public | 9 (6.8%) | 95 (20.3%) | <0.001 |
| Not reported | 0 (0.0%) | 8 (1.7%) | 0.130 |
| Academic | 0 (0.0%) | 5 (1.1%) | 0.233 |
| Other | 2 (1.5%) | 15 (3.2%) | 0.301 |
| **Industry affiliation among authors** |  |  |  |
| At least one author | 98 (74.2%) | 256 (54.7%) | <0.001 |
| First author | 3 (2.3%) | 24 (5.1%) | 0.162 |
| Corresponding author | 4 (3.0%) | 40 (8.6%) | 0.032 |
| **Analysts' affiliation** |  |  |  |
| Only non-industry | 42 (31.8%) | 229 (48.9%) | <0.001 |
| Both by industry and non-industry | 51 (38.6%) | 104 (22.2%) | <0.001 |
| Only by industry analysts | 28 (21.2%) | 97 (20.7%) | 0.903 |
| Not specified | 11 (8.3%) | 38 (8.1%) | 0.937 |
| **Results favored an industry-sponsored intervention** |  |  |  |
| Yes | 102 (77.3%) | 262 (56.0%) | <0.001 |
| No | 8 (6.1%) | 37 (7.9%) | 0.477 |
| No industry-sponsored intervention involved | 22 (16.7%) | 168 (35.9%) | <0.001 |
| Not applicable | 0 (0.0%) | 1 (2.1%) | 0.595 |
| **Data analysis** |  |  |  |
| Analysis code sharing | 6 (4.6%) | 21 (4.5%) | 0.977 |
| Data availability statement | 102 (77.3%) | 376 (80.3%) | 0.439 |
| *Declared data sharing (n =478)* | 88 (86.3%) | 283 (75.3%) | 0.018 |
| **Access to data (n =371)** |  |  |  |
| Request to authors | 17 (19.3%) | 97 (34.3%) | 0.008 |
| Request to repository or archive | 26 (29.6%) | 78 (27.6%) | 0.717 |
| Request to company | 23 (26.1%) | 59 (20.9%) | 0.296 |
| Request to committee, group, or unit | 7 (8.0%) | 22 (7.8%) | 0.956 |
| Access unspecified | 9 (10.2%) | 17 (6.0%) | 0.176 |
| Data available to others | 6 (6.8%) | 10 (3.5%) | 0.185 |
| **Full protocol availability** |  |  |  |
| Published in peer-reviewed journal | 77 (58.3%) | 333 (71.2%) | 0.005 |
| Published online | 22 (16.7%) | 60 (12.8%) | 0.256 |
| None | 33 (25.0%) | 75 (16.0%) | 0.018 |

| **Statistical analysis plan availability** |  |  |  |
| --- | --- | --- | --- |
| Published in peer-reviewed journal | 63 (47.7%) | 291 (62.2%) | 0.003 |
| Published online | 26 (19.7%) | 66 (14.1%) | 0.115 |
| None | 43 (32.6%) | 111 (23.7%) | 0.04 |

<sup>a</sup> Trials included in the original study that do not include any genetic or -omics application. <sup>b</sup> There were 106 studies with combination funding in which at least one of the funders was industry

## Discussion

Our study maps the current landscape of how genetic and other omics-based technologies are utilized in the most cited clinical trials, revealing ongoing patterns and challenges. About a fifth of these impactful trials reported the use of such technologies in these publications, which is a considerable proportion. However, the vast majority of these uses pertained to simple genetic testing, usually focused on specific mutations. Far fewer trials incorporated more comprehensive genomic technologies, despite the rich options that genomic technologies could offer for clinical research ^11^, and other -omics platforms were rarely used. The analyzed trials largely pertained to oncology applications, almost always had some industry involvement, and almost always found favorable results for the sponsor. Some transparency indicators such as public availability of full protocols and of statistical analysis plans tended to be less common in trials using genetic or -omics technologies.

Major challenges in evaluating and using -omics platforms in applied, large-scale clinical research may be related to economic and regulatory factors ^12,13^. Usually only highly funded research can handle the significant costs and navigate the regulatory requirements. Moreover, economic evaluations and cost-effectiveness are key factors that policymakers or decision-makers may consider when introducing a new test or technology for reimbursement. Cost considerations may hinder or even penalize innovative new -omics technologies and influence the allocation of the usually limited research funding. This may lead to a lower prevalence of clinical trials involving - omics technologies ^14,15^.

Precision medicine has made the most strides to-date in oncology ^16^. The focus on cancer may be attributed to the high prevalence of actionable somatic mutations for which targeted therapies are developed. Advancements in genome sequencing technologies ^17^, have made them increasingly cost-effective and capable of processing large volumes in a short time, but most applications in clinical trials focus on single mutations. In most other areas of medicine, personalized approaches are still underdeveloped^18–20^. Additional molecular layers that could provide comprehensive insights into disease mechanisms and therapeutic responses are often overlooked ^21^.

The focus on targeted therapies, which concentrate more on actionable somatic mutations ^22,23^, is a research agenda heavily funded by the industry ^24,25^. Overall 6 out of 7 of the most-cited trials are funded by the industry, and three quarters have at least one author affiliated with an industry sponsor ^26^. For trials involving genetics or other -omics technologies the industry involvement is even higher, and this applies to both oncology and other specialty applications.

This dominance of industry funding raises several questions about potential bias, not only in study design and analysis but also in the choice of clinical outcomes and their reporting and interpretation. Seventy-three percent of industry-funded genetic or other -omics trials reported results favoring the sponsor’s intervention, a proportion significantly higher compared to non-genetic trials. The proportion was even higher in the subset of oncology trials. Concerns that financial interests may influence the outcomes and interpretations of clinical research have also been raised in the past for oncology trials and targeted treatments ^25,27^. Transparency patterns of the genetic and other -omics trials reveal a possibly lower proportion of available protocols and statistical analyses plans, and this may not facilitate transparency. Moreover, even though data availability statements are very common, the presence of data directly available without restrictions remains extremely low.

Our study has some limitations. First, we only focused on the primary publications of each trial, and it is possible that several of these influential trials may also have additional secondary publications^28^. Some of them may deal with genetic or -omics-related secondary analyses or other aspects. Therefore, the penetration of these technologies in these influential trials may be even higher. However, the primary publications are those that carry the most weight in the literature. Second, we cannot extrapolate from our sample to the vast majority of less cited trials. However, our sample is important to study, since it comprises the trials that are shaping the most hot areas of medical research and practice. Third, we did not consider other types of biomarker measurements, e.g., immunocytochemistry, single protein markers or imaging, that are also used occasionally in efforts to personalize medical treatments. Such different types of biomarkers may have different patterns of use and misuse compared to genetic and -omics technologies.

Acknowledging these caveats, our evaluation shows that while genetics has already a substantial role in shaping the agenda of influential clinical research, most of the uses of these technologies in clinical trials remain rudimentary. There is a large window for enhancing the incorporation of newer, more comprehensive technologies. The value of such enhanced applications will need to be rigorously tested, safeguarding the research agenda from sponsor bias and maximizing transparency.

## Data Availability

All data produced in the present study are available upon reasonable request to the authors

## Acknowledgement

The work of Luigi Russo has been supported by the European Network Staff Exchange for Integrating Precision Health in the Healthcare Systems project (Marie Skłodowska-Curie Research and Innovation Staff Exchange no. 823995).

## Data statement

The raw data can be shared by the authors upon request.

